# Epidemiological, phylogenetic, and resistance heterogeneity among *Acinetobacter baumannii* in a major US Deep South healthcare center

**DOI:** 10.1101/2024.03.22.24304710

**Authors:** Emma Graffice, Derek B. Moates, Sixto M. Leal, Megan Amerson-Brown, Juan J. Calix

## Abstract

*Acinetobacter baumannii* (*Ab*) disease in the U.S. is commonly attributed to outbreaks of one or two monophyletic carbapenem resistance (CR) *Ab* lineages that vary by region. However, there is limited knowledge regarding *Ab* epidemiology and population structure in the U.S. Deep South, and few studies compare contemporary CR and carbapenem-susceptible (Cs) *Ab*, despite prevalence of the latter. We performed a 12-year time series analysis of *Ab* cases in a large hospital in Birmingham, AL, and 89 isolates from an ongoing surveillance project started in November 2021 were analyzed by whole genome sequencing and antibiotic susceptibility testing (AST). Cumulative CR rate among 2462 cases since 2011 was 19.4%, with increased rates during winter months resulting from seasonal changes in Cs*Ab* incidence. Sequenced CR*Ab* belonged to clonal complex (CC) 1, CC108, CC250, CC2, and CC499. Most CR*Ab* CCs were comprised of isolates that clustered apart from U.S. counterparts in phylogenetic analysis, despite being identified in unrelated cases occurring ≥3 months apart. In contrast, 38/47 (81%) Cs*Ab* isolates each belonged to a distinct CC. CR*Ab* isolates displayed lineage-distinct AST features, including unique carbapenem resistance genetic determinants and presumptive heteroresistance behaviors unique to CC108 and CC499 isolates. This first comprehensive analysis of *Ab* cases in the U.S. Deep South revealed epidemiological trends consistent with those in other regions and an unusually high degree of phylogenetic diversity among regional CR*Ab* isolates. We also describe emergent U.S. CR*Ab* lineages whose unconventional antimicrobial resistance features must be integrated into ongoing diagnostic, treatment, and surveillance efforts.

## Introduction

The looming global health threat posed by multidrug resistant (MDR) organisms is best exemplified by the ESKAPE pathogens: vancomycin-resistant *Enterococcus faecium*, methicillin-resistant *Staphylococcus aureus*, extended-spectrum beta-lactamase (ESBL)-producing *Klebsiella pneumoniae*, carbapenem-resistant (CR) *Acinetobacter baumannii (Ab*), multidrug-resistant *Pseudomonas aeruginosa*, and ESBL *Enterobacter* species^1^. CR*Ab* in particular is considered a top priority for research by the World Health Organization^2^ and U.S. CDC^3^. This is due to its ability to survive in various environments and its various multidrug resistant (MDR) phenotypes mediated by intrinsic *Acinetobacter*-derived cephalosporinase (ADC) class C β-lactamases and OXA class D β-lactamases, polymorphisms in antibiotic targets, and a variety of other resistance genes -- including extrinsic OXA and non-OXA carbapenamases -- acquired via mobile genetic elements (e.g., plasmids, transposons, etc.). CR*Ab* disease is conventionally linked to hospital-acquired (HA) respiratory, endovascular, wound, and urinary infections mainly attributable to a subset of MDR global lineages within the otherwise genetically diverse species^4–6^.

*Ab* strains can be classified according to one of two multilocus sequence type (MLST) schemes^7^, and to avoid confusion, herein we exclusively use the Pasteur MLST scheme^8^. Single locus variant (SLV) sequence types (STs) can be further grouped into monophyletic lineages, or clonal clusters (CC), named according to the prevailing ST. CR*Ab*-associated lineages CC2, CC1 and CC79 comprised 61%, 5% and 3%, respectively, of the ∼3500 genomes available on NCBI in early 2019^9^. U.S. clinical *Ab* populations have historically reflected global trends, with CC2 comprising 85% of CR*Ab* isolates in the first nationwide survey performed between 2008-2009^10^ and 64% of isolates from CDC sentinel sites between 2013-2017^11^.

In contrast, CC2 comprises <5% of sequenced isolates from Latin American countries^9^, demonstrating that regional population structures can vary significantly. Similarly, observations from U.S. single-center studies reveal wide variation over locations and time^11–15^. For example, most pre-2012 CR*Ab* isolates in a St. Louis, MO healthcare system were CC2 or CC79^10^, but local subclones of the unrelated ST406 (CC406) and ST499 (CC499), the latter of which was first detected in Chicago in 2010^16^, rapidly became the most common CR*Ab* between 2017 and 2019^13^.

Awareness of *Ab* CCs propagating in a region could inform clinical practice, as recent evidence suggests that CR*Ab* lineages each display unique susceptibility patterns to non-carbapenem antibiotics and may differ in their epidemiological behavior^13^. Notably, little is known regarding the population structure of *Ab* in the US Deep South. Other than the inclusion of six Florida isolates^10^, published surveys have not included isolates from Deep South/Southeast region, where unique climate patterns and socioeconomic factors may selectively favor traits in this environmental microbe absent from pools in other regions. In addition, most recent surveys exclude cases associated with carbapenem-susceptible (Cs) *Ab,* despite these being a common cause of *Ab* disease and better representing the genetic diversity of the species. Indeed, 59.3% and 22.0% of CR*Ab* isolates sequenced in the St. Louis study (n=59) belonged to ST499 and ST406 outbreaks, respectively, but each sequenced Cs*Ab* isolate (n=31) belonged to a distinct ST, reflecting a much more sporadic nature of Cs*Ab* infections^13^. Here we report early findings from our investigation of *Ab* population dynamics in the U.S. Deep South.

We compare the epidemiology of Cs*Ab* and CR*Ab* isolates identified over 12 years in a large healthcare system in Birmingham, Alabama and characterize the current population structure of clinically-relevant *Ab* by isolate whole genome sequencing (WGS). We report a link between emergent lineages and atypical antimicrobial resistance phenotypes, and contextualize findings within the greater population structure of *Ab* throughout the U.S.

## Methods

### Study location and period

This study was approved by the University of Alabama at Birmingham (UAB) Institutional Review Board (IRB# 30003572 and 30008212) and was performed in the UAB Healthcare system from July 1, 2010 to December 31, 2022. UAB is a large integrated inpatient and outpatient healthcare system serving Birmingham, Alabama, USA and surrounding areas. It employs Cerner electronic medical record (EMR) systems, which was queried using the Informatics for Integrating Biology and the Bedside (i2b2) tool hosted by the UAB Center for Clinical and Translation Science (CCTS). UAB Hospital and affiliated clinics use a central UAB Clinical Microbiology Laboratory (UAB-CML) housed on the main campus.

### Retrospective study design and definitions

For all cases in which *Acinetobacter* isolates were identified via standard of care culture, we retrieved patient demographics, culture tissue source, hospital day of culture (if applicable), and antibiotic susceptibility testing (AST) interpretation results. The species identity of bacterial isolates was determined by automated biochemical methods or matrix-assisted laser desorption/ionization and time-of-flight mass spectroscopy (MALDI-TOF MS) performed in the UAB-CML. Only the first isolate per patient (“index culture”) reported as following was eligible for inclusion: “*Acinetobacter baumannii*” (n=419), “*Acinetobacter baumannii* complex” (n=651), “*Acinetobacter baumannii* complex/*haemolyticus*” (n=5), or “*Acinetobacter baumannii*/*haemolyticus*” (n=1387). Per UAB-CML protocols, all *Acinetobacter* isolates are routinely tested for susceptibility via the Beckman Coulter Microscan WalkAway plus AST system. Common AST within the UAB-CML includes ampicillin-sulbactam (SAM), cefepime (FEP), ceftazidime (CAZ), ciprofloxacin (CIP), gentamycin (GM), levofloxacin (LVX), trimethoprim-sulfamethoxazole (SXT), tobramycin (TOB), imipenem (IPM), and meropenem (MEM). AST and susceptibility reporting is performed according to Clinical and Laboratory Standards Institute (CLSI) guidelines^17^.

Isolates non-susceptible to either imipenem or meropenem by minimum inhibitory concentration (MIC)-AST were defined as CR. Cases were classified into 5 anatomical categories according to isolate source: “respiratory,” “skin and soft tissue/musculoskeletal” (SST/MSK), “urinary,” “blood” (including isolates obtained from central lines, endovascular devices, or grafts), or “other.” Cases were defined as HA if the index culture was performed ≥48 hours from time of hospitalization and before discharge. All other isolates were defined as nHA. To evaluate for seasonal trends among cases identified in 2011-2022, cases were grouped into four quarters according to the month index culture was obtained: 1 (January-March), 2 (April-June), 3 (July-September) and 4 (October-December).

### Prospective isolate banking

Since November 2021, clinical isolates typed as an *Acinetobacter* species by MALDI-TOF MS in the UAB-CML were identified via biweekly query of the UAB Cerner EMR system and eligible for inclusion. Subclones from original culture plates were transferred to the research laboratory for processing. If more than one morphologically distinct colony on culture plate was identified as *Acinetobacter,* all colonies were stored. For this staging study, we arbitrarily chose 89 presumptive *Ab* isolates identified between November 1, 2021 and November 31, 2022 for sequencing and susceptibility testing analysis (Data S1). Herein, study isolates are named by number (e.g., isolate 002 denotes UAB_Ab002, etc.).

### Whole genome sequencing, assembly and comparative genomics

A full description of the well-established processing pipelines^13^ used for genome assembly and comparative genomics analyses is provided in the Supplementary Text. The earliest isolate with a MLST and/or antimicrobial resistance gene (ARG) profile per person was denoted as an index isolate, with subsequent isolates denoted as non-index isolates.

### Kirby-Bauer AST

Study isolates were tested by Kirby-Bauer disk diffusion AST (KB-AST) in our research lab with Mueller-Hinton agar against sulbactam/ampicillin (SAM), cefepime (FEP), ceftazidime (CAZ), meropenem (MEM), imipenem (IPM), trimethroprim/sulfamethoxazole (SXT), ciprofloxacin (CIP), gentamicin (GM) and doxycycline (DOX). Zone of clearance (ZOC) breakpoints were determined and interpreted according to CLSI guidelines^17^. In cases where subpopulations of distinct colonies were repeatedly observed growing up to the border of the antibiotic disk but readily distinguishable from the lawn’s major border, we subcloned bacteria from colonies at the disk border (“disk subclone”) or from the main lawn (“lawn subclone”). Subclones were plated and incubated at 37°C for 16 hours on LB Miller agar without antibiotics and retested by KB-AST the next day. Plate images were acquired from a height of 6.3 inches using an iPhone 14 Pro Max with 1x lens and arranged using Adobe Illustrator^18^.

### Systematic review of *Ab* isolate population structure in the U.S

Full description of the systematic literature review and comparative genomic meta-analysis is provided in the Supplementary Text. Briefly, a screen on NCBI database was performed on September 18, 2023 and identified 774 genome sequences of *Ab* isolates identified in U.S. populations since 2010 in peer-reviewed reports. We performed *de novo* genome assembly using NCBI SRA files and, when available, extracted epidemiological and microbiological metadata (Data S2). Genomes were binned into CCs according to MLST analysis, and comparative genomic analysis was done within each CC. In the case where a CC of interest was absent among identified genomes (i.e., CC108), we manually screened for genomes on NCBI whose metadata indicated they were isolated in the U.S. since 2010.

### Statistical analyses

Univariate analyses were performed with SPSS v29 (IBM, USA) or RStudio software v 4.1.2^19^. Chi-squared or odd ratio test were performed for comparing categorical variables. Mann-Whitney test with Bonferroni adjustment for multiple comparisons was performed for continuous variables. Statistical significance was defined as *p* value <0.05.

## Results

### Retrospective analysis reveals diverse clinical presentations of *Ab* cases at UAB

We identified 2462 cases associated with *Ab* index cultures at UAB between July 2010 and December 2022, including 478 (19.4%) cases with CR*Ab* (Data S3). Cs*Ab* but not CR*Ab* case occurrence displayed seasonality, with the selective increase in incidence of Cs cases during third calendar quarters (Figure 1A) resulting in decreased CR rates between May and October (Figure 1B). 41.6% (826/1984) of Cs*Ab* cases were HA, compared to 54.2% (259/478) of CR*Ab*. Though CR*Ab* cases were more likely than Cs*Ab* to be HA (54.2% versus 41.6%, OR [CI95%]=1.66 [1.36-2.03]), HA rates fluctuated over time among CR*Ab*. HA rates were 76.9% and 85.7% in late 2010 and early 2011, respectively, but were 27.4% and 45.5% in both halves of 2022 (Data S3). This trend coincided with CR*Ab* cases in later periods being identified earlier during hospitalization (Figure 1C). In contrast, Cs*Ab* HA rates and median hospital day of isolation were similar in all study periods (Figure 1D).

**Figure 1.**
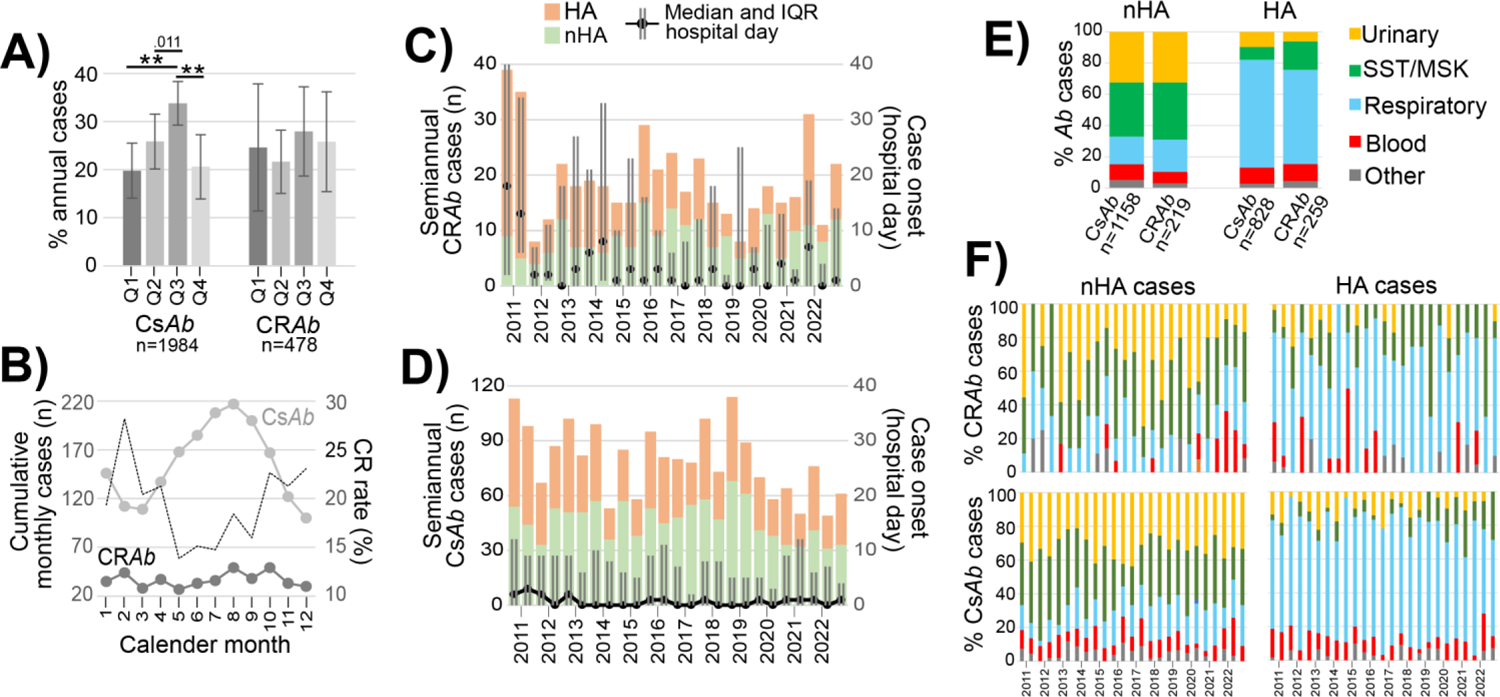
Divergent epidemiological features of *Ab* cases at UAB, July 2010-December 2022. A) Proportion of annual carbapenem-susceptible *A. baumannii* (CsA*b*) and carbapenem-resistant Ab (CR*Ab*) cases occurring in each calendar quarter (Q1-Q4) during 2011-2022. **, <0.001. B) Cumulative case total for each month (left y-axis) of Cs*Ab* and CR*Ab* (light and dark gray, respectively) and monthly CR rate (dotted line, right y-axis) among cases from 2011-2022. C,D) Semiannual totals (left y-axis) of hospital-acquired (HA) and non-hospital acquired (nHA) CR*Ab* (panel C) and Cs*Ab* (panel D) cases. Lines and error bars depict the median and interquartile range (IQR) for hospital day of isolation among cases in each period (right y-axis). Panel C key corresponds to both panels. E) Percentage of CR*Ab* and Cs*Ab* index isolate from each tissue sources among nHA (left) and HA (right) cases. F) Percentage of index isolates from each source among CR (top row) and Cs (bottom row) cases in each study period, separated by nHA (left column) and HA (right column).

Cs*Ab* and CR*Ab* displayed comparable distribution of isolate tissue sources when stratified according to onset (Figure 1E), i.e., respiratory isolates predominated among HA cases, while urinary and SST/MSK isolates predominated among nHA cases in nearly all study periods (Figure 1F). Lastly, CR*Ab* was more likely to be isolated from males (OR [CI95%] = 1.49 [1.21-1.84]), and females comprised less than 40% of Cs*Ab* respiratory cases (37.2%) and CR*Ab* respiratory (37.7%), SST/MSK (33.1%) urinary (34.7%), and other (31.6%) cases (Table S1). There was no difference in average patient age between Cs*Ab* (49.0 years) and CR*Ab* (50.3 years, p=0.178) cases. In summary, CR*Ab* and Cs*Ab* were both implicated in various case types at UAB, albeit with differential features regarding seasonality, timing of index culture, and patient sex distribution.

### 2021-2022 UAB *Ab* isolates demonstrate phylogenetic heterogeneity

To describe the *Ab* population structure in our cohort, we performed WGS of 89 *Ab* isolates identified in 2021-2022, including 73 index isolates from all tissue sources, obtained from 68 case patients (Figure 2, Data S1). We identified 31 assigned STs (including ST2139 and ST2140, which were newly assigned in this study) and 11 unassigned STs distributed among 39 CCs (Table S2). Ten cases had >1 *Ab* isolate included in our WGS analysis, and three patients had two index isolates from unrelated STs (Figure S1). 58.9% (n=43) of index isolates belonged to nine CCs, each containing ≥2 index isolates obtained from unrelated cases occurring ≥30 days apart (Figure 2, Table S2). Of these, CC108, CC164 and CC427 each contained isolates of two STs. The remaining index isolates (n=30) each belonged to unrelated STs (Data S4).

**Figure 2:**
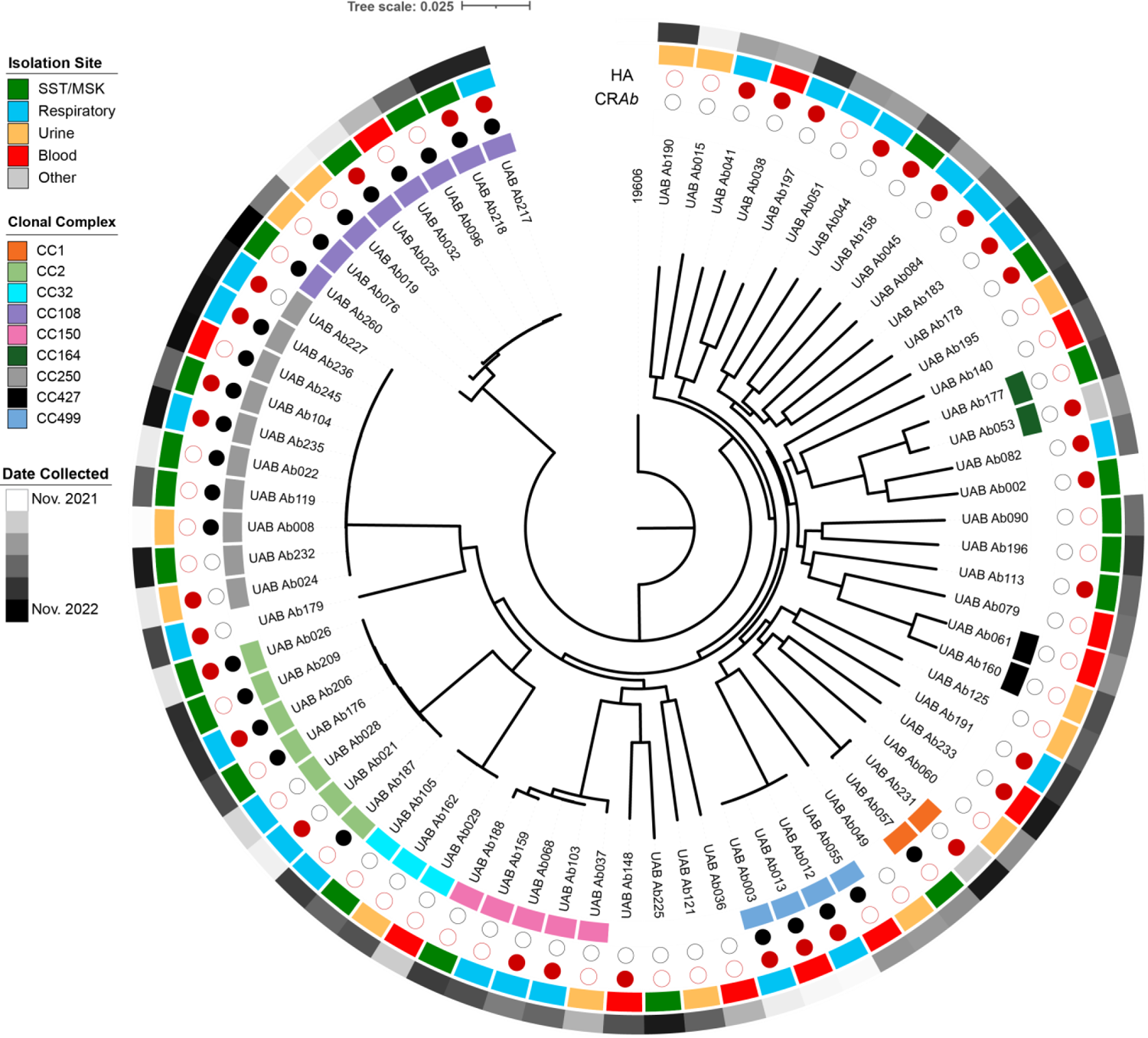
Diversity of *Ab* isolates at UAB hospitals, 2021-2022. Maximum likelihood phylogenetic tree of UAB *Ab* index isolates according to alignment of 2072 core genes, rooted to the genome of reference strains 19606. Inner label ring denotes CC with >1 index isolate. Filled circles denote CR*Ab* and isolates associated with HA cases. Outer rings represent isolate metadata according to the corresponding keys, as listed in Data S1.

### Antimicrobial susceptibility was linked to lineage dependent and independent genetic elements

We investigated whether UAB CR*Ab* lineages display distinctive AST profiles, as observed in recent studies^13^. Our cohort was comprised of 26 CR*Ab* and 47 Cs*Ab* index isolates (Figure 3, Data S1). CC108 and CC499 were exclusively comprised of CR*Ab*, CC150, CC164, CC32 and CC427 were comprised exclusively of Cs*Ab*, and CC1, CC2 and CC250 were comprised of both. All remaining isolates were Cs*Ab*. ARG analysis revealed all CC499 isolates encode OXA-24, the CC108 isolate 260 encodes OXA-499 (an OXA-143-like carbapenemase), and OXA-23 is encoded in nearly all other CR isolates except isolate 032 (Figure 3). Along with six other CC108 isolates, isolate 032 encodes the FtsI A515V polymorphism, a penicillin-binding protein 3 mutation predicted to decrease carbapenem binding to its active site^20^. In summary, carbapenem resistance wholly correlated with the presence of attributable genetic elements (Figure 3, Data S4).

**Figure 3:**
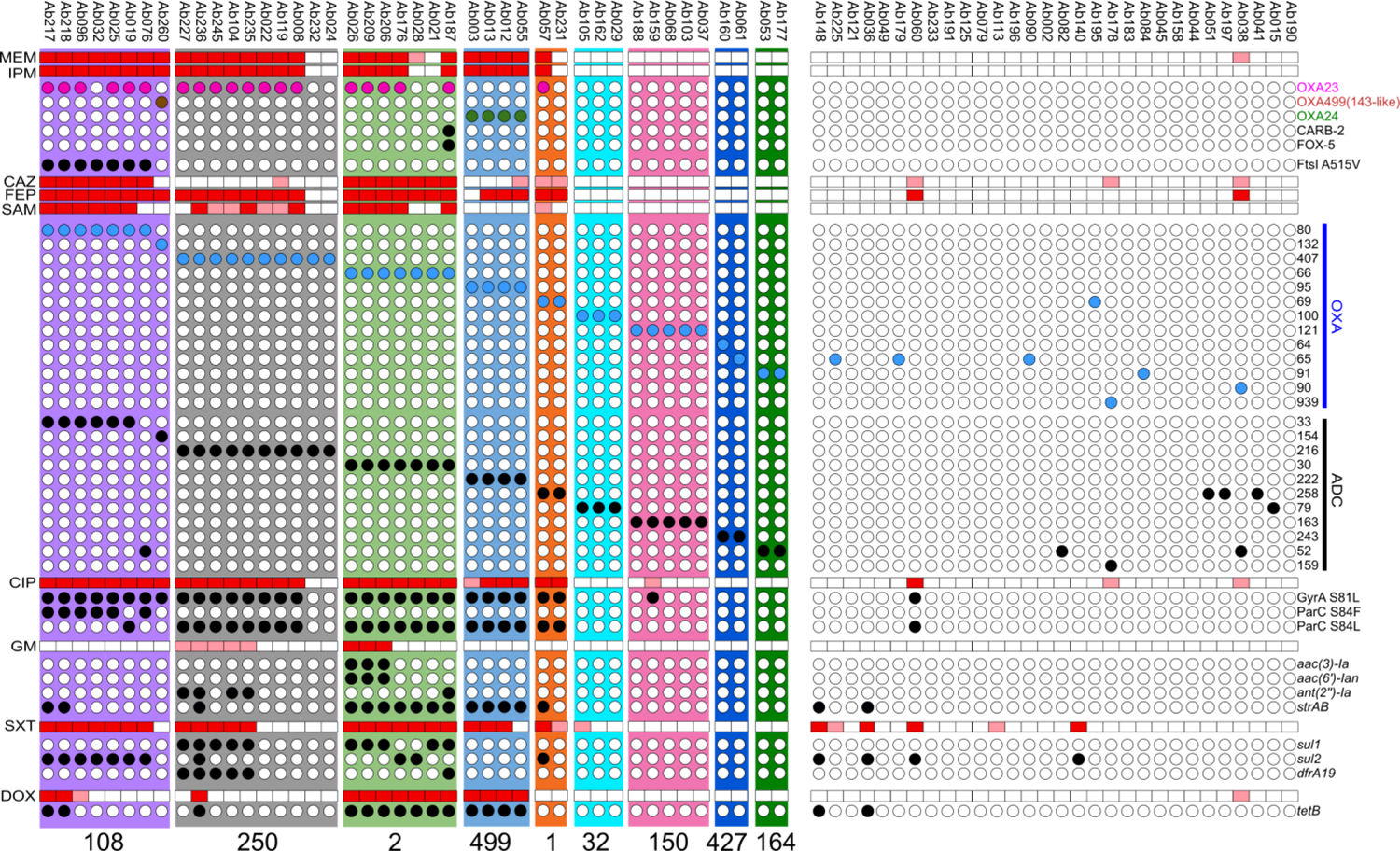
Antibiotic susceptibility and resistance genetic element content of UAB index isolates (top). Tested antibiotics are listed on left and grouped by antibiotic class. Results are presented as “resistant” (red), “intermediate” (pink), or “susceptible” (white) according to CLSI guideline interpretation of Kirby-Bauer results. Circles below each AST group denotes presence (fill circle) or absence of associated resistance genetic elements listed on right. Intrinsic OXA and ADC alleles are listed by allele number. For clarity, only genetic elements present in isolates belonging to multi-isolate CCs (colored columns with labels below) are included. Results for all UAB isolates and genetic elements are listed in Data S4.

The identity of intrinsic OXA and ADC alleles was largely conserved within a lineage and rarely shared between lineages (Figure 3). Independent of OXA-23 presence, CC1 and CC2 isolates displayed greater FEP and CAZ resistance than other Cs*Ab*, including the CC250 Cs*Ab* isolates (Figure S2). Conversely, CC250 and CC499 CR*Ab* isolates displayed CAZ susceptibility indistinguishable from CC150 and other Cs*Ab*. CC108 isolates displayed the lowest detectable levels of susceptibility across all beta-lactam antibiotics in KB-AST (Figure S2). We observed comparably greater intra-CC variation in the susceptibility to other antibiotic classes, which strongly correlated with the presence of resistance elements repeatedly shared by genomes in multiple lineages (Figure 3 and Figure S2). The only major discrepancy was SXT non-susceptibility in CC499 isolate who lacked the *sul1/sul2* gene putatively conferring resistance in all other lineages, which is a trait observed among CC499 isolates in prior studies^13^.

### Atypical resistance phenotypes displayed by CC108 and CC499 isolates

Results from clinical laboratory AST by microbroth dilution (MB-AST) generally matched Kirby-Bauer AST (KB-AST) for most isolates, but two consistent discrepancies were noted (Data S1, S5). First, CC108 isolates displaying the lowest detectable susceptibility to SAM in KB-AST were reported as “intermediate” or “susceptible” according to MB-AST. Second, despite CC499 isolates being reported as “susceptible” to SXT according to MB-AST and lacking conventional resistance gene elements observed in other SXT-resistant isolates, they displayed decreased susceptibility to SXT in KB-AST. Closer review of KB-AST plates found that CC108 isolates against IPM and SAM and CC499 isolates against SXT each displayed subpopulations extending between their main lawn margin and the antibiotic disk borders (Figure 4A). Subclones obtained from the lawn or colonies at the antibiotic disk border were incubated overnight without antibiotics, and secondary KB-AST was performed (Figure 4B). Each subclone pair yielded similar results in secondary KB-AST (Figure 4C-E). Similar SXT-related phenomenon was observed when testing ST499 isolates identified prior to 2019 in St. Louis (Figure S3)^13^. No other tested isolates displayed similar patterns in KB-AST.

**Figure 4:**
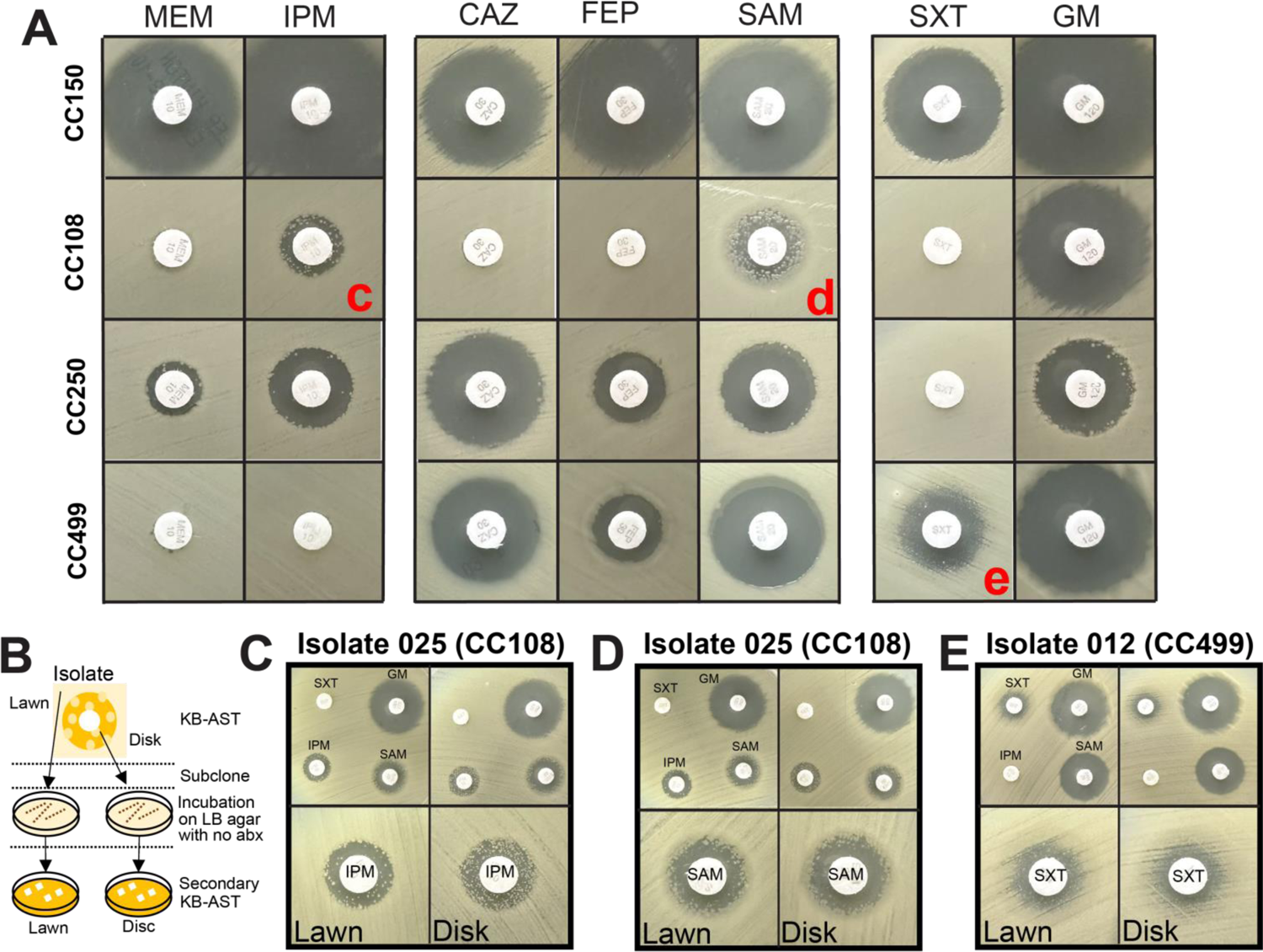
KB-AST identifies susceptibility-variable subpopulations associated with CR*Ab* lineage and antibiotic type. A) Representative pictures of the most commonly observed KB-AST results for main CCs in our cohort (left), against different antibiotics (top). Red letters denote samples that were subcloned and incubated overnight on LB agar with no antibiotic followed by secondary KB-AST, as depicted in panel B. C-E) Secondary KB-AST of “lawn” (left) and “disk” (right) subclones. Photos show results from multiple antibiotics (top) and magnification of zones of clearance around antibiotics of interest (bottom). MEM, meropenem; IPM, imipenem; CAZ, ceftazidime; FEP, cefepime; SAM, ampicillin-sulbactam; SXT, trimethoprim/sulfamethoxazole; GM, gentamicin.

### Meta-analysis of contemporary U.S. *Ab* population structures

The equal presence of multiple unrelated CR*Ab* lineages in our cohort was unexpected. To understand how strains in our cohort relate to their lineages in the greater U.S. *Ab* population, we integrated 645 *Ab* genomes from strains isolated in U.S. since 2010 that passed QC (Data S2) and 545 genomes were used for CC-specific comparative analyses (Table S2). UAB CC2 isolates occupied three branches within two previously described clades^14,21^, one branch in Clade C (isolates 021 and 187) and two in Clade A (isolate 028 and all the others, respectively) (Figure 5). Most CC1, CC499, CC108 and CC250 UAB isolates occupied exclusive branches within their complexes, though two CC250 (isolates 024 and 232) and CC108 isolates (isolates 076 and 260) were on distinct branches.

**Figure 5:**
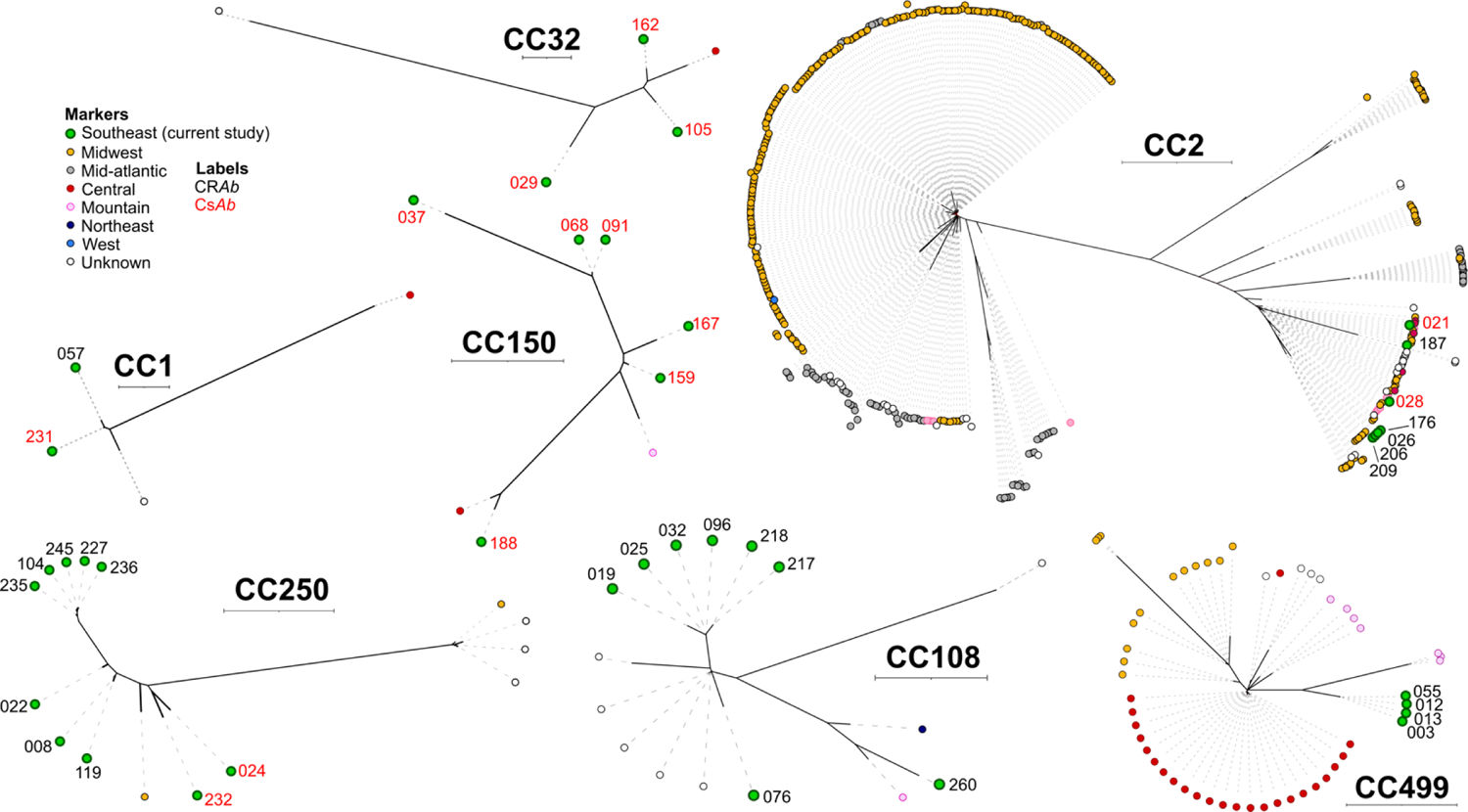
Intra-CC phylogeny of U.S. *Ab* populations identified since 2010. Unrooted maximum likelihood phylogenetic trees drawn according to alignment of core genes shared by isolates within each CC. Tree scales located under CC labels represent to 0.001 substitutions per site, with CC1 and CC32 trees presented as half scale compared to others. Leaf markers represent the regions where each isolate was identified, per the inset key. Numbered labels denote study number of carbapenem-resistant (black) or -susceptible (red) isolates in the UAB study.

Conversely, UAB CC150 and CC32 genomes were broadly distributed within their respective trees. No CC427 or CC164 isolates were identified in the U.S. meta-analysis, and two of the most common U.S. lineages, CC406 (also known as USA-clone-1^22^) and CC79 (also known as Clade E^14^), were absent from the UAB cohort (Table S2).

In ARG analysis of the total U.S. cohort there was within-lineage variability regarding strains harboring resistance genetic elements for non-beta-lactam antibiotics (Data S2). In contrast, the allelic identity of intrinsic OXA and ADC genes facilitated the delineation of phylogenetic subclades within clonal complexes, most of which included isolates obtained in different studies and regions (Figure S4). The vast majority of isolates encoding a carbapenemase contained OXA-23. However, OXA-24/72 was the prevalent carbapanemase among CC2 isolates in subclades A2, C2 and G (Figure S4B), half of CC79 isolates, two unrelated subclades in CC406, and nearly all non-Mountain region CC499 isolates (Figure S4C). Notably, the FtsI A515V polymorphism was encoded by isolates only in CC2 subclade B2 (Figure S4B) and one CC108 subclade that included isolates lacking any OXA carbapenamase (Figure S4C). CC406 was the only CR*Ab*-associated complex in which most isolates had no identifiable carbapenem resistance element. CC150 and CC32 isolates did not encode carbapenamases (Figure S4D).

## Discussion

We report a comprehensive analysis of clinical cases associated with *Ab* isolates in a major Deep South U.S. medical center. Aiming to uncover the microbiological landscape of *Ab* propagating in our region, we followed an ecological study design to characterize the presentation of *Ab-*associated cases and the population structure of clinical isolates. Investigation of treatment strategies, outcomes, and transmission networks spurring their presence is the subject of ongoing study in our regional cohort.

Few recent surveys include both Cs*Ab* and CR*Ab* in their analyses despite the former being more prevalent among *Ab* clinical isolates in many regions^6,23^. CR*Ab* cases were more likely to be HA in our cohort, though the trend of cases being identified earlier in a hospital course did not coincide with a decrease in carbapenem resistance (Data S3), and tissue source distributions of CS and CR isolates were indistinguishable (Figure 1E-F). The observed link between tissue source distribution and case onset as well as the presence of seasonality only among Cs*Ab* (Figure 1A-B) is consistent with prior reports^6,24^. Altogether, these findings support that our cohort is comparable to *Ab* in other U.S. medical centers.

Genomic analysis identified multiple clusters comprised of isolates collected ≥90 days apart, from different tissue sources, and associated with HA and nHA cases (Figure 2), and many of these isolates belong to subclades unique to our region. Attributing *Ab* clusters to hyperlocalized outbreaks would require a formal clonality analysis beyond the scope of our current study. Regardless, these observations imply the existence of contemporary microbial pools persisting in the region and outside our hospital, as shown in other U.S. regions^13,14^. While 26 index CR*Ab* belonged to five CCs, 47 Cs*Ab* isolates belonged to 37 unrelated lineages. The identification of multiple cases in a single U.S. center caused by single lineages (i.e. CC150 and CC32) has not been reported. However, compared to CR*Ab* isolates, there tended to be greater distance between UAB Cs*Ab* isolates within their respective CCs (Figure 5). The relatively decreased relatedness among Cs*Ab* isolates is consistent with recent reports^13^, and support that Cs*Ab* cases are result of sporadic cross-over events from environmental sources.

As a species living in undefined environmental niches, *Ab* harbors a very diverse genetic pool even compared to other Gram-negative pathogens^25,26^. Though historically most CR*Ab* disease has been attributed to CC2 isolates, unrelated lineages have recently emerged in the U.S. For example, the first reported incidence of ST499 isolation occurred in 2010^16^, but CC499 has now become the second most common lineage among U.S. CR*Ab* isolates and the dominant lineage in some regions^11,13^. CC108 had been only sporadically observed in prior studies, but our finding support it is capable of establishing a stable presence in a region. Lastly, ST250 was first identified in a 2007 isolate from Puerto Rico^27^, rarely identified in prior U.S. surveillance, but repeatedly isolated from unrelated cases over the course of our study. As reflected by the relative diversity of CR*Ab* lineages concurrently propagating in our region, climate in the U.S. Deep South may be more permissive to *Ab* phylogenetic diversity than other sites where surveillance has previously been performed^11^, and we predict similarly diverse *Ab* population structures would be revealed upon investigation of other Deep South sites. These findings highlight that we should avoid generalizing historical or even multicenter study findings to local *Ab* population structures, as the lineages responsible for local cases can differ drastically from site to site.

The value of surveilling local population structures remains to be determined, as the contribution of *Ab* phylogeny to pathogenesis and clinical outcomes is unclear. However, consistent with prior findings^28^, we observed that CR*Ab* lineages can harbor unique antimicrobial resistance features that may influence local medical practices. For example, OXA-23 has historically been the most prevalent carbapenamase among CR*Ab* and, thus, is a target of current and emergent rapid molecular detection tools in clinical labs^29^. However, we identified U.S. isolates belonging to CC2, CC108, and CC499 that display carbapenem resistance via other resistance determinants (i.e., OXA-24 or the FtsI A515V polymorphism), and the mechanism of carbapenem resistance among CC406 isolates remains to be identified^13,22^ (Figure S4). During the preparation of this manuscript, Sabour, et al. reported that carbapenemase gene content among CR*Ab* differed by region, and these genes did not appear to always confer the same degree of resistance^28^. Though Sabour et al. did not evaluate whether degree of carbapenem resistance was dependent on lineage, lineage-associated differences were observed previously^13^ and in our study (i.e., OXA-23-associated IPM resistance was greater among CC2 isolates compared to CC250 isolates, Figure S2). Altogether, this implies that the utility of OXA-23-based testing varies according to which U.S. lineages are circulating in the region.

Another concerning finding was the discordance between MB-AST and KB-AST for SAM against CC108 isolates and for SXT against CC499 isolates. The repeated occurrence of subpopulations with different susceptibilities and the instability of increased resistance phenotype in subclones together suggest the presence of heteroresistance in these isolates, though further work is required to confirm and characterize this. Why these traits are not readily identifiable via MB-AST, and how the spontaneous occurrence of increased resistance against sulbactam and trimethoprim/sulfamethoxazole influences clinical outcomes or susceptibility to new combination agents (i.e., sulbactam-durlobactam), must be evaluated. The dynamic nature of these resistance phenotypes is reminiscent of phase variation, but the mechanisms through which the phenotype is achieved require further study. This is especially true for CC499, which appears able to achieve elevated resistance to SXT without harboring any of the genetic elements typically associated with SXT resistance in other *Ab* lineages (i.e. *sul1* or *sul2,* Figure 3).

Though consistent with recent reports from other U.S. locations^11–13^, our study was performed in a single healthcare center and the generalizability of findings to other global *Ab* populations remains to be determined. Importantly, our work begins to show how knowledge of isolate lineage could influence clinical decision-making and sets a foundation to explore how lineage may influence other aspects of *Ab* disease, such as virulence, clinical prognosis, and propensity to be spread among certain patient populations.

## Data Availability

All raw sequence files and assemblies derived from UAB study strains are available under NCBI BioProject PRJNA1005294. All other data in the present study are available upon reasonable request to the corresponding author.

---

**Figure S1:**
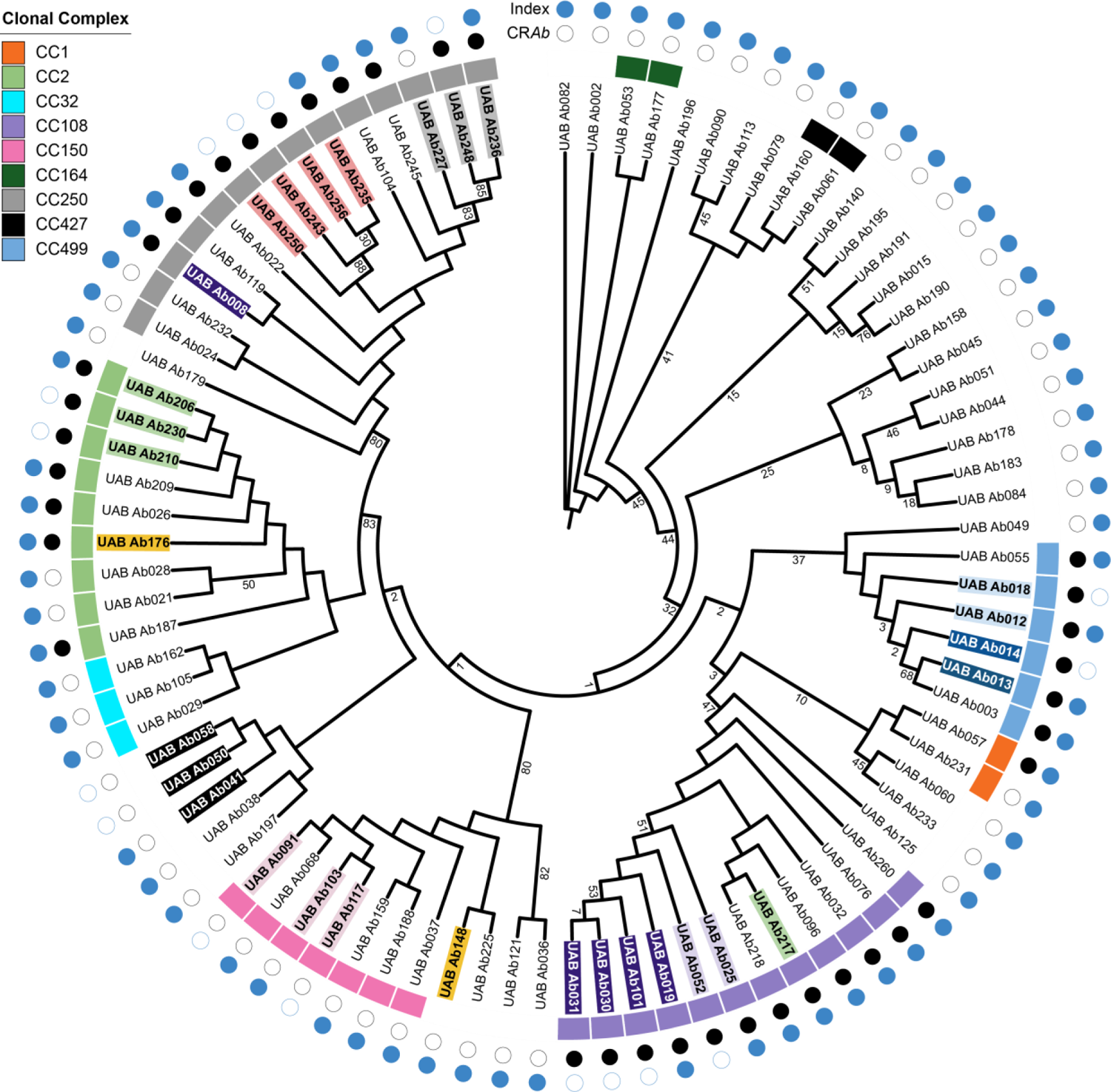
Phylogenetic clusters among all *Ab* sequenced isolates in this study. Maximum likelihood tree of all UAB *Ab* isolates, rooted to midpoint. Branches with bootstrap values <90% are labeled. Highlighted and bolded leaves represent isolates obtained from the same patient, matched according to color. Outer boxes are color coded according to key to denote CCs containing >1 index isolate. Carbapenem resistance and index isolates are denoted by filled black and blue circles, respectively.

**Figure S2:**
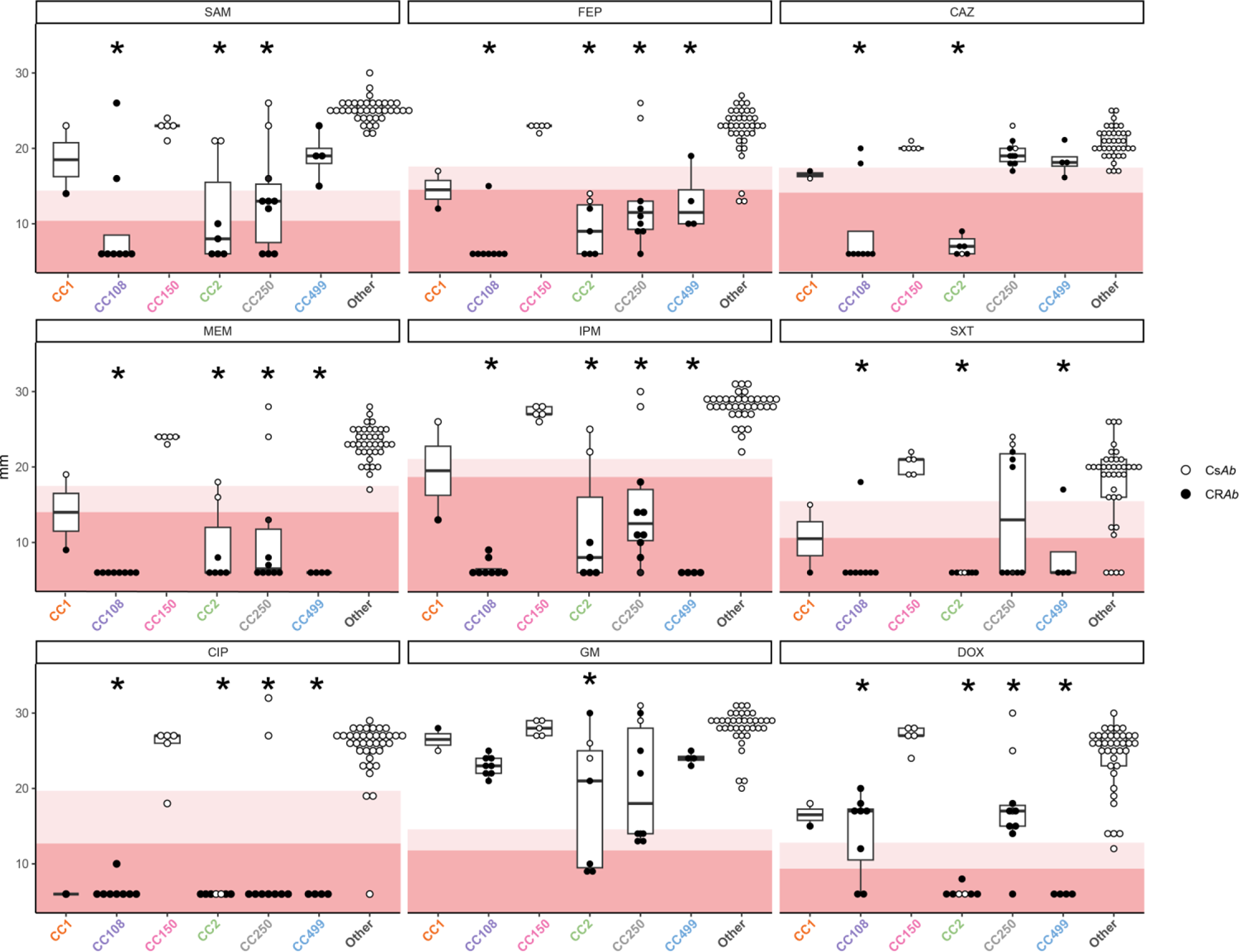
Kirby-Bauer antimicrobial susceptibility testing (KB-AST) of UAB isolates. Antibiotic susceptibility, according to zone of clearance (mm, y-axis), of CR*Ab* (black dots) and CsAb (white dots) index isolates grouped by CC. Backgrounds highlight ranges for “resistant” (dark) and “intermediate” (light) susceptibility, per CLSI guideline interpretation. Box-plot center lines denote medians; box limits, upper and lower quartile values and whiskers, 1.5 times the IQR. Medians were compared to CC150 using Mann-Whitney test with Bonferroni adjustment for multiple comparisons, with asterisks denoting P < 0.05. Abbreviations: CAZ, ceftazidime; CIP, ciprofloxacin; DOX, doxycycline; FEP, cefepime; GM, gentamicin; IPM, imipenem; MEM, meropenem; SAM, ampicillin-sulbactam; SXT, trimethoprim-sulfamethaxazole.

**Figure S3:**
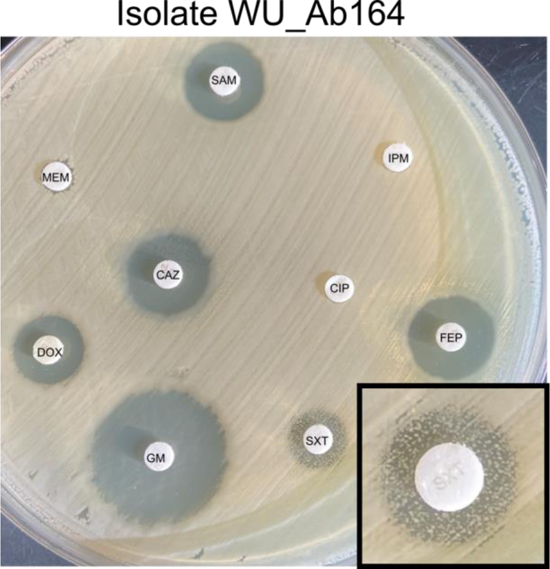
CC499 isolates from an unrelated cohort also display variable SXT susceptibility. **Photos of** KB-AST results of CC499 isolate WU_Ab164, against of all tested antibiotics and magnification of zone of clearance around SXT disk (inset). Abbreviations: CAZ, ceftazidime; CIP, ciprofloxacin; DOX, doxycycline; FEP, cefepime; GM, gentamicin; IPM, imipenem; MEM, meropenem; SAM, ampicillin-sulbactam; SXT, trimethoprim-sulfamethaxazole.

**Figure S4:**
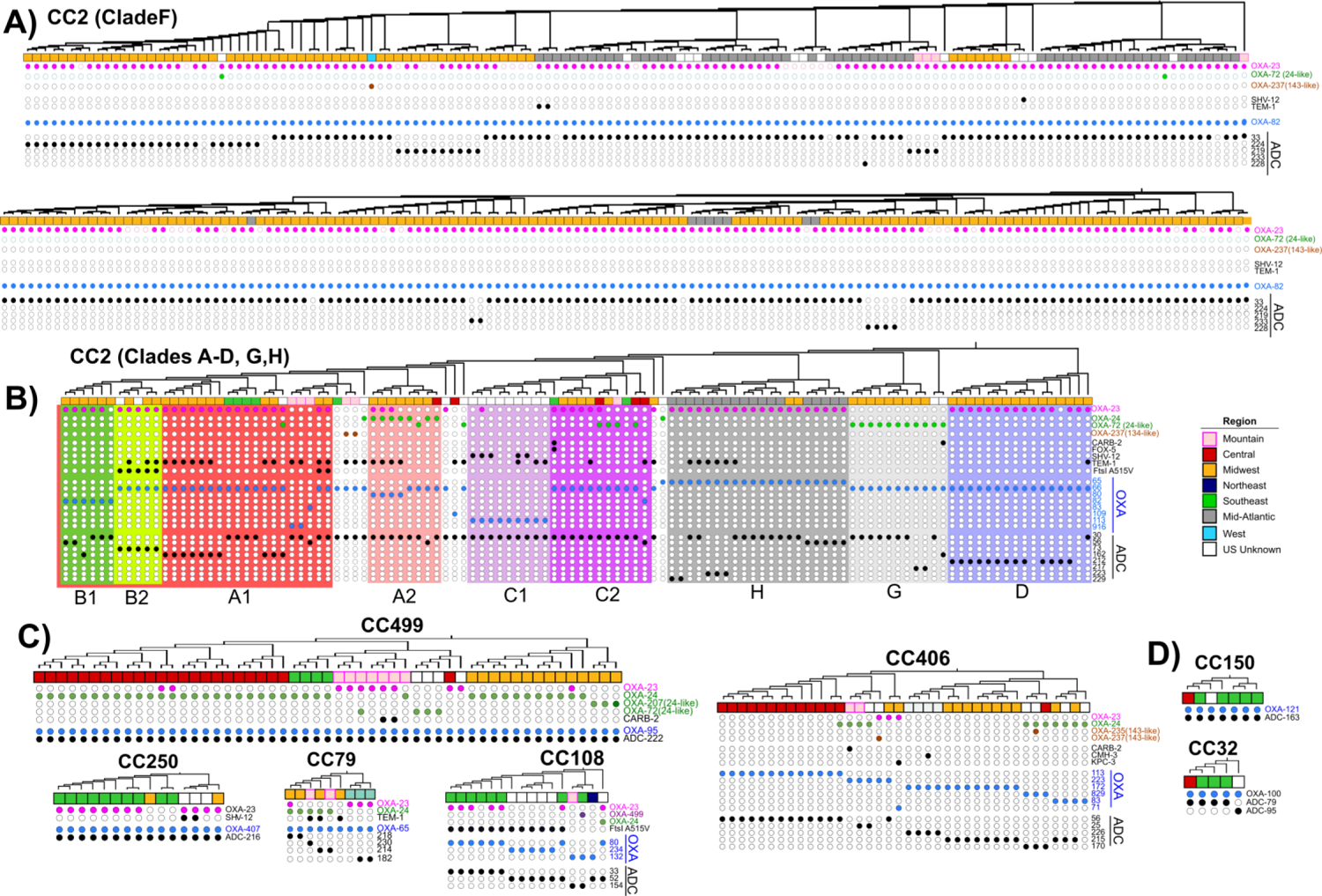
CC-wide ARG content among major U.S. lineages associated with CR*Ab* (panels A-C) and CS*Ab* (panel D). Squares denote an isolate’s region with color key in panel B. Top rows of matrices in panels A-C show OXA carbapenamases (labels colored according to subclass), followed by non-OXA genetic elements conferring β-lactam resistance in black. The lower lines denote intrinsic OXA (blue) and ADC (black) lactamase alleles. All genomes contain an ADC allele, but ADC alleles that were detected in only one genome are omitted for clarity.

**Table S1:** Distribution of patient sex among UAB *Ab* cases.

|  | Carbapenem susceptible *A. baumannii* specimen type |  |  |  |  |  | Carbapenem resistant *A. baumannii* specimen type |  |  |  |  |  |
| --- | --- | --- | --- | --- | --- | --- | --- | --- | --- | --- | --- | --- |
|  | Blood | Other | Respiratory | SST/MSK | Urine | Total | Blood | Other | Respiratory | SST/MSK | Urine | Total |
| Male | 110 | 39 | 483 | 241 | 231 | 1104 | 19 | 13 | 130 | 85 | 65 | 311 |
| Female | 91 | 45 | 292 | 228 | 224 | 880 | 25 | 6 | 71 | 42 | 22 | 166 |
| % female | 45.3% | 53.6% | 37.7% | 48.6% | 49.2% | 44.4% | 56.8% | 31.6% | 35.3% | 33.1% | 25.3% | 34.7% |
| Sum | 201 | 84 | 775 | 469 | 455 | 1984 | 44 | 19 | 201 | 127 | 87 | 478 |

SST/MSK, skin, soft tissue and musculoskeletal

**Table S2:** Intra-lineage phenotypic and genotypic breakdowns.

| Clonal complex | UAB Cohort |  |  |  |  |  |  |  |  |  |  |  |  |  | U.S. Cohort (NCBI) |  |  |  |  |  |
| --- | --- | --- | --- | --- | --- | --- | --- | --- | --- | --- | --- | --- | --- | --- | --- | --- | --- | --- | --- | --- |
|  |  |  |  |  |  | Tissue sources (n) |  |  |  |  | Genomic analysis |  |  |  |  |  |  |  |  |  |
|  | Days collected | Pasteur MLST | Index isolates | n CRAB (%) | nHA (%) | SST/MSK | Urine | Resp. | Blood | Other | Total genes | Core genes | Core genome SNPs | SNP/core gene | Genomes analyzed | Pasteur MLST | Total genes | Core genes | Core genome SNPs | SNPs/core gene |
| CC1 | 167 | 734 | 2 | 1 (50) | 1 (50) | 2 | 0 | 0 | 0 | 0 | 2792 | 3497 | 489 | 0.1398 | 4 | 1, 734 | 4210 | 3250 | 19806 | 6.0942 |
| CC2 | 257 | 2 | 7 | 5 (71) | 4 (57) | 2 | 0 | 5 | 0 | 0 | 4570 | 3183 | 1650 | 0.5184 | 398 | 2, 195, 632, 1573 | 31307 | 2411 | 25625 | 10.6284 |
| CC32 | 175 | 32 | 3 | 0 (0) | 0 (0) | 1 | 1 | 0 | 1 | 0 | 3933 | 3275 | 1878 | 0.5734 | 5 | 32 | 4343 | 3190 | 3231 | 1.0129 |
| CC108 | 340 | 108, 2140 | 8 | 8 (100) | 5 (63) | 3 | 2 | 2 | 1 | 0 | 3861 | 3213 | 3339 | 1.0392 | 16 | 108, 2140 | 4550 | 2600 | 15102 | 5.8085 |
| CC150 | 167 | 150 | 5 | 0 (0) | 3 (60) | 2 | 1 | 2 | 0 | 0 | 4152 | 3360 | 14860 | 4.4226 | 7 | 150 | 4458 | 3265 | 14325 | 4.3874 |
| CC164 | 109 | 164, 1131 | 2 | 0 (0) | 1 (50) | 1 | 0 | 0 | 0 | 1 | 3857 | 3231 | 11631 | 3.5998 | NA | NA | NA | NA | NA | NA |
| CC250 | 344 | 250 | 12 | 10 (83) | 5 (42) | 4 | 2 | 5 | 1 | 0 | 4819 | 3334 | 709 | 0.2127 | 15 | 250 | 4645 | 3140 | 1463 | 0.4659 |
| CC427 | 89 | 427, 848 | 2 | 0 (0) | 0 (0) | 0 | 1 | 0 | 1 | 0 | 4261 | 3094 | 15695 | 5.0727 | NA | NA | NA | NA | NA | NA |
| CC499 | 126 | 2139 | 4 | 4 (100) | 1 (25) | 0 | 0 | 2 | 2 | 0 | 3722 | 3677 | 26 | 0.0071 | 53 | 499, 1180, 2139 | 5034 | 3065 | 12489 | 4.0747 |
| CC406 | NA | NA | NA | NA | NA | NA | NA | NA | NA | NA | NA | NA | NA | NA | 38 | 406, 1088, 1091, 1562 | 6248 | 2641 | 24797 | 9.3892 |
| CC79 | NA | NA | NA | NA | NA | NA | NA | NA | NA | NA | NA | NA | NA | NA | 9 | 79 | 4639 | 3106 | 7655 | 2.4646 |
| NA indicates where an analysis could not be performed. |  |  |  |  |  |  |  |  |  |  |  |  |  |  |  |  |  |  |  |  |

